# Quantifying the number of people who would benefit from HIV pre-exposure prophylaxis (PrEP) in the United States: a comparison of behavioral, acquisition-risk based, and economic metrics

**DOI:** 10.1101/2025.09.29.25336900

**Authors:** Minttu M. Rönn, Athena P. Kourtis, Yizhi Liang, Lijia Zheng, Teresa Puente, Ya-Lin A. Huang, Weiming Zhu, Rupa R. Patel, Jeffrey Wiener, Karen W. Hoover, Michelle Van Handel, Nicolas A Menzies, Joshua A Salomon

**Author notes:** Corresponding Author: Minttu M Rönn.

## Abstract

**Purpose:** We developed metrics to estimate the number of people who could benefit from PrEP using clinical, behavioral, and economic considerations.

**Methods:** We estimated the distribution of annual HIV acquisition risk in the U.S. population and the number who would benefit from PrEP based on HIV acquisition risk thresholds. Estimates were generated for men who have sex with men (MSM), men who have sex with women (MSW), women who have sex with men (WSM), and people who inject drugs (PWID). Populations were stratified by state, age, and race and ethnicity. Adult PWID were stratified by state and sex. We also derived a measure anchored on a willingness-to-pay threshold to gain one quality-adjusted life year (QALY).

**Results:** We estimated 31–57% of MSM could benefit from PrEP by HIV acquisition risk thresholds, and 30% when using the cost-per-QALY threshold. For PWID, estimates ranged from 7% (cost-per-QALY) to 60% (highest risk threshold). MSW and WSM had the lowest proportions estimated to benefit (0-11%), but the absolute number of individuals remained large due to the size of these populations.

**Discussion:** These estimates provide a broader framework in which to examine need for PrEP at the population and program level in the United States.

## Introduction

Pre-exposure prophylaxis (PrEP) is highly effective in preventing HIV acquisition.^1–3^ The U.S. Preventive Services Task Force^4^ and Centers for Disease Control and Prevention (CDC)^5^ recommend PrEP for HIV prevention. However, implementation and scale-up remain challenging. Overall uptake of PrEP remains below targets, with disparities relative to the burden of HIV^6,7^.

Behavioral indications for PrEP are based on sexual– and injection-drug-use-related acquisition risk.^8,9^ For sexual acquisition, PrEP is indicated for people who, in the past six months, had a diagnosed bacterial sexually transmitted infection (STI), had more than one sexual partner with unknown HIV status and inconsistent condom use, or a sexual partner living with HIV with a detectable viral load. Among people who inject drugs (PWID), PrEP is indicated for those who have injected drugs and shared injection equipment in the past six months.^8,9^ An estimated 31,800 new HIV infections occurred in 2022 in the United States.^10^ HIV acquisition risks vary across transmission risk groups, with higher incidence rates among men who have sex with men (MSM) and PWID; 67% of all incident infections occurred among MSM.^10^ Acquisition varies by geography, age and race/ethnicity.^10^

Smith and colleagues developed an approach to estimate numbers of persons who could benefit from PrEP combining behavioral indications and HIV risk indicators.^11^ The method was used by the CDC to report PrEP coverage between 2018 to 2023 (“former CDC estimate”). The method is anchored in the proportion of MSM with behavioral indications for PrEP using data from the National Health and Nutrition Examination Survey (NHANES). For other groups, the number who could benefit from PrEP is estimated using the distribution of MSM and non-MSM populations in state-level HIV diagnosis data. This applies behavioral indications for PrEP in NHANES for MSM and scales estimates for other groups according to the relative HIV diagnosis rates between groups. The approach does not account for the vastly different population sizes of the different HIV transmission risk groups.

We developed metrics based on behavioral indications, acquisition risks, and economic considerations to estimate who could benefit from PrEP. These reflect differing data sources, assumptions, and foundational premises for estimation. These estimates are programmatic in nature, guiding public health interventions and resource distribution. They can also identify key uncertainties and data gaps to guide future research.

## Methods

### Populations with behavioral indications for PrEP

We used NHANES^12^ data to estimate the proportion of the adult population with behavioral indications for PrEP based on their self-reported sexual behavior, by HIV transmission risk group (MSM, men who have sex with women [MSW], women who have sex with men [WSM]) and age group (18-24, 25-34, 35-44, 45-54, 55+ years). For MSW and WSM, behavioral indications are defined as people who reported more than one sexual partner in the past year, and who either used condoms inconsistently or reported having been diagnosed with gonorrhea, which approximates clinical guidance for PrEP.^5^ Smith *et al*. estimated 24.7% of MSM to have indications for PrEP, based on the same indications as for MSW and WSM, in addition to including chlamydia diagnosis as an indication, based on CDC PrEP guidelines. However, other behavioral surveys for MSM have estimated up to 51% of MSM reporting indications for PrEP.^13^ MSM with fewer behavioral indications may remain at increased risk of HIV acquisition. Therefore we considered a broader definition: having at least one anal intercourse partner and inconsistent condom use, or a diagnosis of gonorrhea or chlamydia in the past year, which accounts for 50.6% of the MSM population. We used this definition to determine the proportion of MSM with behavioral indicators.

We estimated the population size of sexually active adolescents aged 13–17, using data from the Youth Risk Behavioral Surveillance^14^ and National Survey of Family Growth^15^ from respondents aged 15-17 years (Supplementary Material). In the absence of nationally representative data, we assumed that among sexually active adolescents, the proportions with behavioral indications for PrEP were the same as in the youngest group of sexually active adults, aged 18-24. We used population size estimates stratified by state, age, race and ethnicity, and sex from the American Community Survey (ACS microdata, 2019 5-year sample) to estimate numbers of people with behavioral indications for PrEP via sexual exposure.^16^ To estimate population sizes of MSM, we used published state-level estimates of the percentage of men who are MSM, as reported in Grey *et al*.^17^

To estimate behavioral indications for PrEP among people who inject drugs (PWID), we used estimates from a meta-analysis of the population size of people who are currently injecting drugs.^18^ We included the adult population of men and women aged 18-44 years by U.S. state to generate the population size of PWID with behavioral indications for PrEP,^16^ combining estimates of current PWID with estimates of the proportion of PWID who report sharing injection equipment.^19^ In the absence of more detailed data, proportions varied by sex, but were assumed to be constant by state, age and race/ethnicity. The more limited age range was selected to more accurately reflect the population size of current PWID. For all populations, when we report people with behavioral indications for PrEP, we use estimates from a calibrated HIV acquisition risk model, described below, to compare estimates that are internally consistent.

### HIV acquisition risk model

We modeled HIV acquisition risks using a mechanistic Bernoulli process modeling framework, as used in other estimates of HIV acquisition risks.^3,20^ The model estimates the average annual HIV acquisition risk among a subgroup of individuals with behavioral indications for PrEP. We developed separate models for sexual acquisition risk and injection drug use acquisition risk, and we assumed these risks were additive among those susceptible to both. For sexual acquisition risk, the population was stratified into subgroups defined by state, transmission risk group (MSM, MSW, WSM), age group (13-24, 25-34, 35-44, 45-54, 55 years), race and ethnicity (non-Hispanic White [White], non-Hispanic Black [Black], Hispanic, non-Hispanic Multiracial [Multiracial], non-Hispanic Native Hawaiian or Other Pacific Islander [NHOPI], and non-Hispanic American Indian or Alaska Native [AIAN]).

The sexual acquisition risk model estimates annual HIV incidence as a function of number of sex partners, and number of sex acts and condom use per partnership. The parameter estimates, by transmission risk group and age, were derived from NHANES from respondents with behavioral indications for PrEP (survey cycles between 2009-2016; further details in the Supplementary Material). NHANES data are nationally representative, and parameter estimates derived from NHANES were assumed to apply to all states. HIV acquisition risk also depends on the transmission probability by act (specific to transmission risk group), condom use efficacy by act, prevalence of HIV in the partner pool, and viral suppression among people with HIV in the partner pool. Prevalence of HIV and viral suppression in the partner pool were informed by HIV surveillance data at state level by transmission risk group.^21^ These two measures accounted for the heterogeneity in HIV risk between states. The model included relative risks by race and ethnicity and transmission risk group,^22^ assumed to be constant by state.

For injection drug use acquisition risk, we used a similar, but simplified, model structure given scarcity of data. Subgroups were defined by state and sex, and the Bernoulli process model quantified risk as a function of number of injection partners, and number of injection events in which a needle was shared. Behavioral parameters were assumed to be constant by state. To parameterize HIV prevalence in the PWID partner pool, we used 3-year cumulative HIV diagnoses among PWID, instead of people with HIV acquired via injection drug use, to approximate the fast turnover in the PWID population (driven by initiation, cessation and relapse of injection drug use),^23^ and HIV acquisition risk from injection drug use was possible only between PWID actively injecting.

We calibrated the HIV risk model to estimates of HIV incidence in 2022^21^ using a Bayesian estimation approach.^24^ We assumed all new HIV infections occurred in populations with behavioral indications for PrEP as defined in this study. HIV incidence estimates were available at the national level, by transmission risk group, age, age among MSM, race and ethnicity, and sex among PWID. Overall incidence at state level was reported for a subset of states, and we calibrated to the state’s incidence when it was reported, and to a pooled residual incidence estimate for all states without state-specific estimates.

We defined prior distributions for key model input parameters (proportion with behavioral indications for PrEP, proportion of PWID, behavioral input parameters for sexual and injection related risks, HIV prevalence and viral suppression in the partner pool, and disparities in HIV acquisition risk by race/ethnicity). Posterior distributions were characterized using a sampling importance resampling.^24^ We sampled 3 million draws from the prior distributions and calculated the corresponding likelihood by draw. We resampled 1000 simulation draws with replacement, with the sampling weight for a given draw proportional to the likelihood estimate for that draw.

### HIV acquisition thresholds approach

The calibrated model predicts the annual average HIV incidence in people with behavioral indications for PrEP. To reflect how HIV infections are distributed in the total population, we plotted Lorenz curves that relate the cumulative estimated new HIV infections over subpopulations ranked by infection rates, to cumulative population size. We can define different thresholds based on what proportion of a given subpopulation should be covered by PrEP if we wanted to avert a certain percentage of total new HIV infections: 100% of HIV infections (total population with behavioral indications for PrEP), 90% of HIV infections (subpopulations who comprise 90% of HIV infections, capturing the subpopulations with the highest incidence) and 75% of HIV infections (subpopulations who comprise 75% of HIV infections, capturing the subpopulations with the highest incidence).

### Cost-effectiveness threshold approach

The second approach defined the number of people who could benefit from PrEP based on a cost-effectiveness criterion that assumed PrEP would be used only where its expected benefits could be attained at less than $100,000 per quality-adjusted life years (QALY) gained. This accounted for the annual cost of PrEP use ($12,376),^25,26^ lifetime costs associated with HIV infection ($478,142),^25,27^ and 4.45 QALYs lost per HIV infection, as reported in Wang *et al*..^25^ This corresponds to 75 persons using PrEP to avert one infection given a willingness to pay threshold of $100,000 per QALY gained. Multiplying this number with the estimated number of incident cases nationally in 2022^21^, by transmission risk group, provides an estimate of the number of people who could benefit from PrEP at a cost-effectiveness ratio below a threshold often used to define an acceptable limit for value for money in economic evaluations.^28^ To derive state-specific estimates in this approach, we distributed the national PrEP quota across states in proportion to transmission risk group-specific HIV diagnoses.

### Smith et al. method (former CDC estimate)

For comparison, we replicated an established method developed by Smith and colleagues to define PrEP need using 2022 HIV diagnosis data.^11^ In the HIV risk model, we defined behavioral indications for PrEP as age-group dependent but, when replicating the Smith et al. approach, we used 24.7% as the proportion of adult MSM with behavioral indications for PrEP across states.^11^ At the state level, equivalence between PrEP ‘need’ and diagnosis distribution is assumed between MSM and non-MSM populations such that PrEP need in any non-MSM population is calculated based on PrEP need in MSM times the ratio of HIV diagnoses in non-MSM populations relative to HIV diagnoses in MSM.

### Outcomes

We report the mean and 95% uncertainty interval from the calibrated HIV acquisition risk threshold model. We report the average annual HIV incidence in a subpopulation, number of estimated HIV infections by subpopulation and percentage of individuals who could benefit from PrEP by estimation approach, with the total US population used as the denominator.

Analytic code is at: https://github.com/mintturonn/prep_need

## Results

The calibrated HIV acquisition risk model replicated the overall trends in estimated incidence in the United States in 2022. Calibration figures are in the supplementary material (Figures S2-S8). Most new HIV infections were estimated to occur in populations with the highest HIV incidence rates, which represent a small proportion of the total population, illustrated in the Lorenz curve showing a highly unequal distribution of infections within the population with indications for PrEP (Figure 1). Lorenz curves for each population suggest varying levels of concentration of HIV across different risk populations, with the most unequal distribution among MSW and WSM (Supplementary Material, Figure S9).

**Figure 1.**
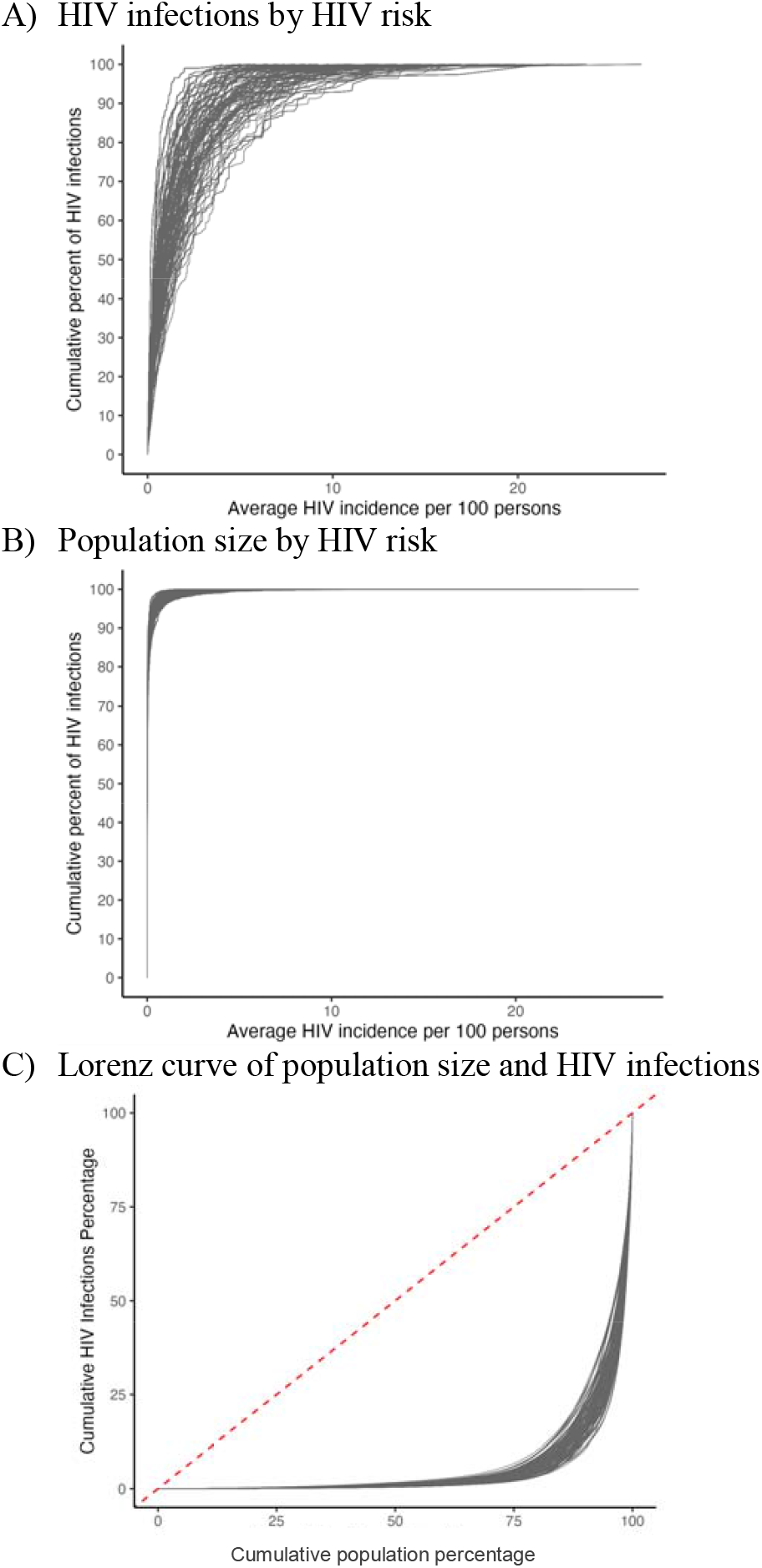
HIV acquisition risk model calibration results. <u>Panel A</u> shows the cumulative proportion (y-axis) of HIV infections by the average annual HIV acquisition risk per 100 persons (x-axis). <u>Panel B</u> shows the cumulative proportion of the population size (y-axis) by the average annual HIV acquisition risk per 100 persons (x-axis). <u>Panel C</u> compares these two cumulative measures using a Lorenz curve (red line represents equality). Each line represents a simulation draw from the calibrated model.

The cost-effectiveness threshold approach estimated 30% of MSM would benefit from PrEP under a limit of $100,000 per QALY compared to 24.7% in the former CDC estimate. In the HIV acquisition risk threshold approach, high proportions of MSM were estimated to benefit from PrEP, ranging from 31% (27%-36%) to 57% (56%-59%) assuming coverage of 75% or 100% of all HIV infections, respectively (Figure 2, Supplementary Material Table S3). Larger differences in estimates were observed for PWID, with 7% estimated to benefit based on cost-effectiveness criteria, but between 26% (0%-44%) and 60% (52%-55%) estimated to benefit based on acquisition risk estimates covering 75% or 100% of infections, respectively.

**Figure 2.**
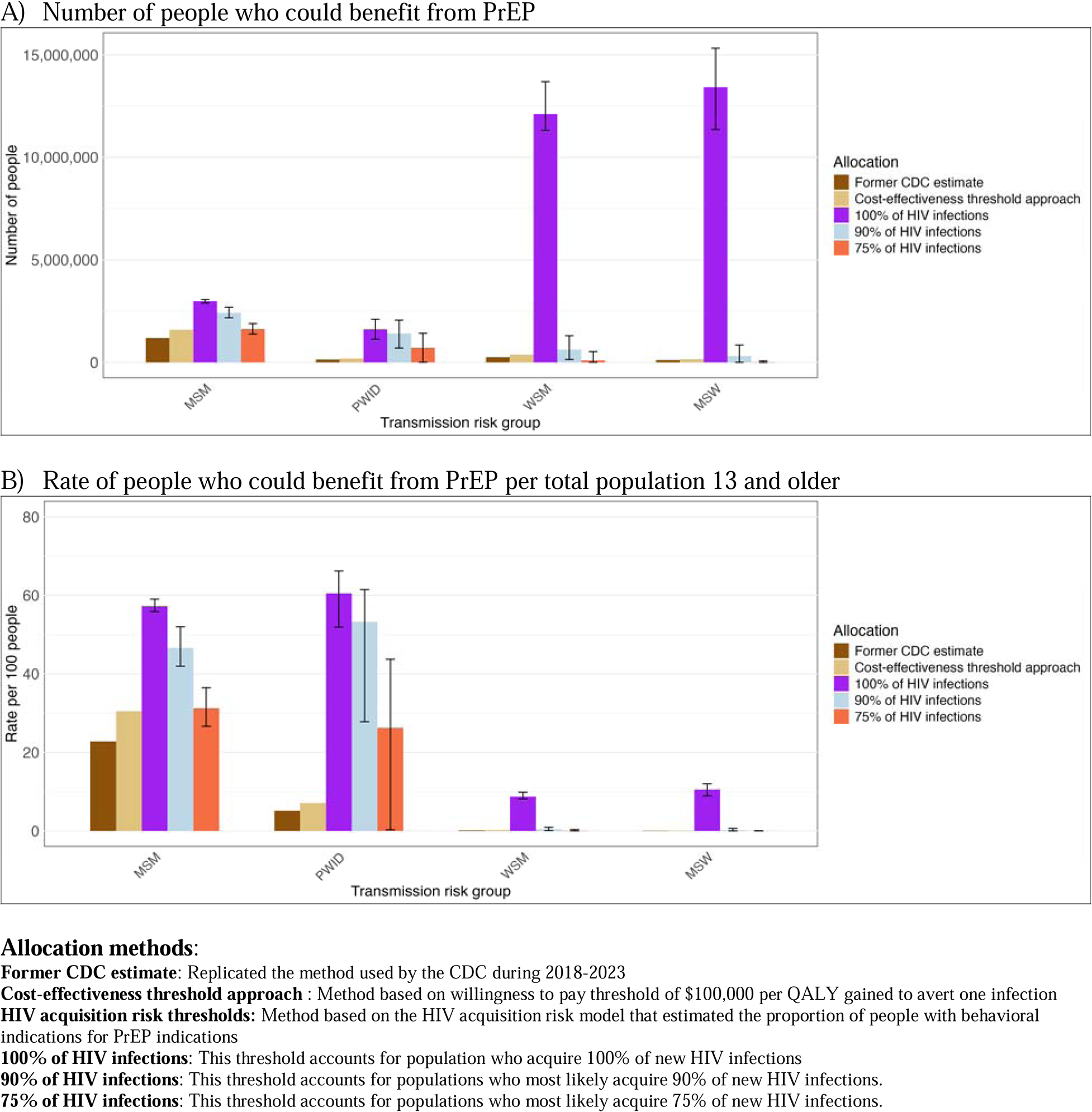
Estimated numbers of people who could benefit from PrEP based on different methods and criteria used, results shown *by transmission risk group*. <u>Panel A</u> displays the number of people, and <u>Panel B</u> displays the rate per 100 persons who could benefit from PrEP per total population 13 years and older.

Among MSW and WSM, 11% (9%-12%) and 9% (8%-10%) were estimated to benefit from PrEP under 100% coverage of HIV infections based on the population with behavioral indications irrespective of underlying HIV risk. Estimated proportions were much lower in all methods that covered less than 100% of HIV infections. Given large population sizes, these still reflect a large absolute number of individuals. For WSM, an estimated 382,000 in cost-effectiveness threshold approach and 99,600 (6,590-529,00) with 75% coverage of HIV infections would benefit from PrEP. For MSW, the estimates were 150,000 and 13,500 (0-74,300), respectively (Figures 2-5; Supplementary Tables S5-S8). Among MSW and WSM, the populations who could most benefit from PrEP were Black WSM 4% (1-7%) and Black MSW 2% (0-6%) with 90% coverage of HIV infections (Figure 4, Supplementary Table S7). For MSM, high proportions of MSM across race and ethnicity groups were estimated to benefit from PrEP in all but the most stringent HIV threshold. With 75% coverage of HIV infections, Black and Hispanic MSM would most benefit from PrEP (54% and 47%, respectively, compared to a 22-33% range for the means estimated for other groups; Table S7).

**Figure 3.**
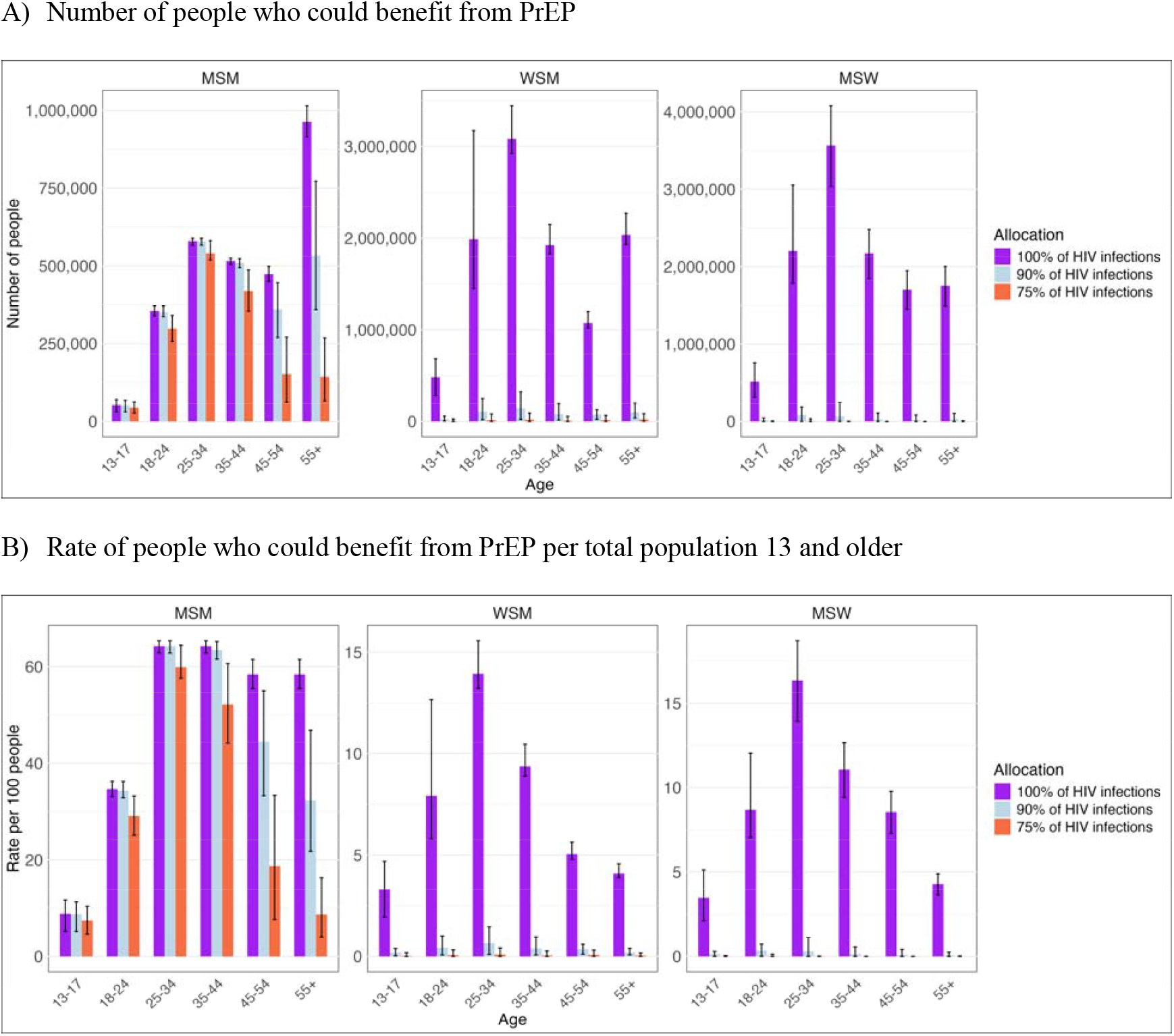
People who could benefit from PrEP based on different HIV acquisition risk thresholds used *by transmission risk group and age*, excluding PWID risk model estimates. <u>Panel A</u> displays the number of people, and <u>Panel B</u> displays the rate per 100 persons who could benefit from PrEP per total population 13 years and older. Y-axes vary between panels.

**Figure 4.**
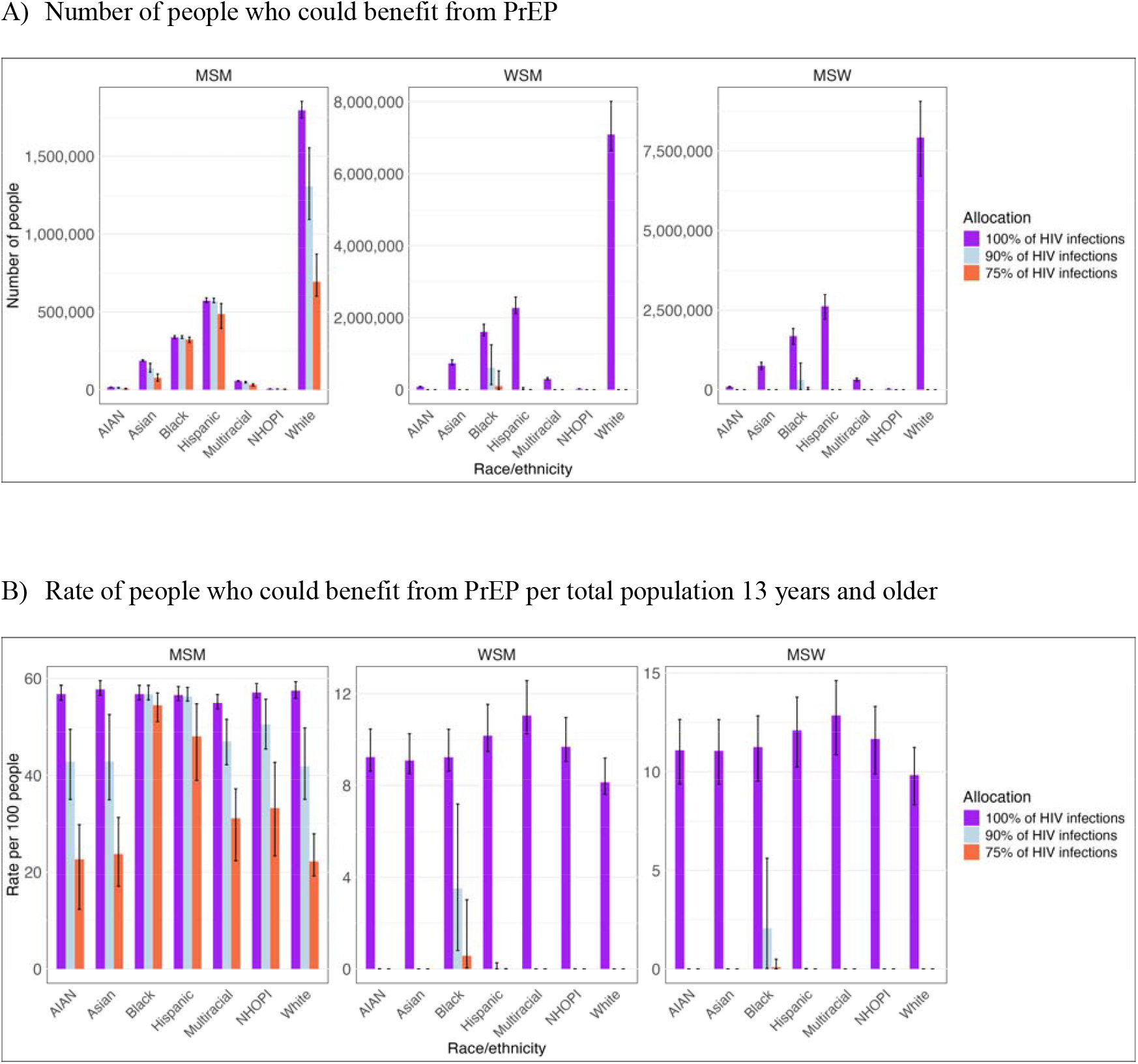
People who could benefit from PrEP based on different HIV acquisition risk thresholds used *by transmission risk group and race/ethnicity*, excluding PWID risk model estimates. <u>Panel A</u> displays the number of people, and <u>Panel B</u> displays the rate per 100 persons who could benefit from PrEP per total population 13 years and older. Y-axes vary between panels.

The population aged 13-17 years was estimated to have a lower proportion who would benefit from PrEP reflecting the lower level of sexual activity in this age group. Among the population aged 13-17 years, MSM had the largest proportion who could benefit from PrEP; 9% (5-11%) and 7% (5-10%) under 100% and 75% HIV infection thresholds, respectively. For MSW and WSM, an estimated 3% (2-5%) of those aged 13-17 could benefit from PrEP under the 100% HIV infection threshold.

Across states, in comparison to the previous CDC estimate and the cost-effectiveness threshold approach, the 75% of HIV infections threshold resulted in similar or lower estimates, while the 90% of HIV infections threshold resulted in similar or higher estimates of numbers who would benefit from PrEP. In all states, estimates based on covering 100% of HIV infections differed substantially from less inclusive estimates, driven mostly by the large number of people with heterosexual exposure (Supplementary Material, Figure S10).

## Discussion

This study explored the impact of different metrics on estimates of the population size that could benefit from PrEP in the United States. These estimates provide a framework in which to examine need for PrEP at the population and program level. The different approaches vary in terms of their data requirements and underlying assumptions. Each method offers unique insights but also presents inherent limitations. MSM remain the population with the highest risk of HIV acquisition, reflected consistently across the different approaches. On the other hand, the estimates varied more substantially for the two largest populations, MSW and WSM. Ultimately, use of one approach over another should reflect both data accessibility and the decision-making context for PrEP implementation.

MSW and WSM overall were estimated to have a small proportion of individuals who would benefit from PrEP. Given the large population size of people with heterosexual exposure and heterogeneities not explored in this study, further research with more granular data is needed to identify those who would most benefit from PrEP in these populations.

All the HIV acquisition risk thresholds identified the population size of PWID who would benefit from PrEP as higher than in the other approaches. There is high prevalence of needle-sharing in this population, and high acquisition risk if HIV is present in the needle-sharing network. While PWID contribute to approximately 7% of incident HIV infections, they only comprise 1.5% of the adult population. Limitations in data for PWID present a particular challenge for PrEP estimation, and HIV epidemiology is associated with outbreaks in PWID communities.

Recently, Kourtis *et al*.^29^ developed a new method to estimate population PrEP need in the United States using HIV incidence estimates to define number needed to treat with PrEP to prevent one additional HIV infection. This method simplifies data requirements but is based on some assumptions related to the threshold used to define who would benefit from PrEP and the proportion of individuals at elevated HIV acquisition risk within each transmission group. This method estimated there were 2.2 million people who would benefit from PrEP. This is broadly like the estimates generated in our cost-effectiveness and 75% of HIV infections threshold approaches.

These metrics share several limitations, most notably the timeliness and availability of data, as they require maintaining complex surveys and deriving incidence estimates based on data on HIV diagnoses. All estimates are subject to evolving epidemiology of HIV, public health practices, and the introduction of new PrEP delivery models. Surveillance measures, such as HIV diagnoses are updated annually. Conversely, the last publicly available data on sexual behavior in NHANES is from cycles collected 2009-2016. Another data limitation relates to estimating benefits of PrEP for 13-17 years olds whose HIV acquisition risk remains less well characterized than that of adults. We did not incorporate time-variant factors into our analysis. Using multiple metrics to estimate PrEP need can offer more comprehensive insights than relying on a single metric and emphasizes the strengths and limitations of different approaches.

## Data Availability

Analytic code is at: https://github.com/mintturonn/prep_need

## Disclaimer

The findings and conclusions in this report are those of the authors and do not necessarily represent the views of the Centers for Disease Control and Prevention, other funding bodies, or the authors’ affiliated institutions.

## Acknowledgments

We would like to acknowledge Cindy Lyles and Paul Farnham for their advice and helpful comments during this study

