## Supplemental Material for "Quantifying the number of people who would benefit from HIV pre-exposure prophylaxis (PrEP) in the United States: a comparison of behavioral, acquisition-risk based, and economic metrics"

---

1. HIV acquisition risk model ... 2
   1.1. Population definitions ... 2
   1.2. Risk equation model ... 5
   1.3. Calibration ... 6
2. Cost-effectiveness threshold approach ... 10
   2.1. Equation ... 10
   2.2. Data ... 10
3. Results ... 11
   3.1. Calibration results ... 11
   3.2. HIV risk model estimates by transmission risk group ... 16
   3.3. Tabulations of number who could benefit from PrEP by allocation criteria ... 17
   3.4. State-level results ... 22
4. References ... 24

### 1. HIV acquisition risk model

#### 1.1. Population definitions

##### 1.1.1. People with PrEP indications

<u>Adolescents, aged 13-17 years</u>
We combined data from National Survey of Family Growth (NSFG) and Youth Risk Behavior Surveillance (YRBS), which measure sexual orientation and having ever had sex. These data were used to determine plausible ranges of sexually active adolescents in population aged 15-17 (Figure S1).

Adolescent men who identified as gay or bisexual were defined as MSM, and adolescent men and women who identified as straight were identified as MSW and WSM, respectively. The higher end of the prior distribution is based on the higher estimate for aged 15-17 (in either NSFG or YRBS). The lower estimate is based on the lowest estimate available for those aged 15 (in either of the data) to acknowledge there would not be as many as many sexually active people in the 13-17 year old population as among 15-17 year olds. The proportion sexually active were similar between the populations, and in the absence of further data by population group, we define a prior distribution range defined where the 95% range set at 10-35% (Table S1). Among the sexually active individuals aged 13-17, we assumed that their behaviors and risk were the same as those aged 18-24, with the same proportion with behavioral indications for PrEP.

**Figure S1.** Proportion of sexually active adolescents (ever had sex) reported in the National Survey of Family Growth (NSFG) and Youth Risk Behavior Surveillance (YRBS)*

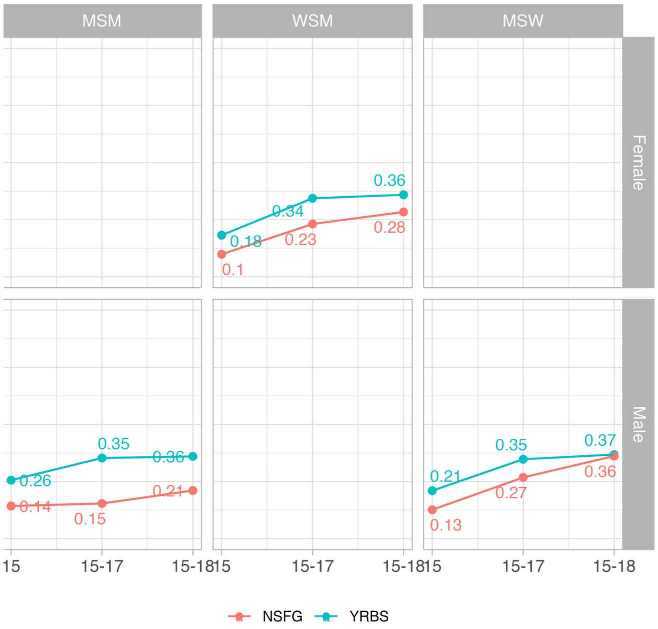

*) We only considered 15, and 15-17 age groups in defined the prior distribution for out parameter. 15-18 is shown for reference only.

Adults, aged 18+

We combined NHANES 4 2-year cycles: 2009-2010 and 2011-2012, 2013-2014, and 2015-2016. We used the survey package in R to derive estimates accounting for the survey design in variance estimation. We restricted all population to data to HIV negative population, and the variable is from the laboratory sample, and the survey weight was constructed from the laboratory survey weight (MEC exam weight). Variance was estimated using the logit method (using svyby and svyciprop functions in the survey package).

MSM: State-specific estimates of the proportion of men who have had a male partner in the past year was used.[1] We assumed this proportion remained constant across age groups. MSM were considered to have PrEP indications if they were not living with HIV, had >1 male sex partner in the past 12 months, and reported inconsistent condom use or had chlamydial, or gonococcal infection. Proportion with PrEP indications was analyzed by age group. For MSM, with smaller population size in NHANES, we defined one range for ages 18-44 and one range for MSM aged 45+ .

MSM population definition

- In the past 12 months, with how many males have you had anal or oral sex? At least one man reported to be included as MSM. All subsequent questions restricted to MSM

Sexual activity

- In the past 12 months, with how many men have you had anal sex? Binary variable constructed, 1 if reported at least 1, 0 if reported 0

Condom use

- In the past 12 months, about how often have you had vaginal or anal sex without using a condom? Binary variable constructed, 1 if reported 'never' or 'about half the time', 0, if reported 'always', 'less than half the time', or 'not always but more than half the time'

STI

- In the past 12 months, has a doctor or other health care professional told you that you had chlamydia?
- In the past 12 months, has a doctor or other health care professional told you that you had gonorrhea, sometimes called GC or clap? Binary variable constructed, 1 if reported yes to either, 0 if report no to both.

MSW: We used NHANES[2] to estimate age-specific proportion of men if they were not living with HIV and they had >1 female sex partner in the past 12 months, and reported inconsistent condom use or had gonorrhea infection. Proportion with PrEP indications was defined by age group, and we enforced the observed age pattern by defining the PrEP indications in the youngest age group, and the other age groups were defined as a function of the youngest age group.

Sexual activity

- In the past 12 months, with how many women have you had any kind of sex? Binary variable constructed, 1 if reported at least 1, 0 if reported 0
- In the past 12 months, with how many males have you had anal or oral sex? Reported none to be included as MSW

Condom use

- In the past 12 months, about how often have you had vaginal or anal sex without using a condom? Binary variable constructed, 1 if reported ‘never’ or ‘about half the time’, 0, if reported ‘always’, ‘less than half the time’, or ‘not always but more than half the time’

STI
- In the past 12 months, has a doctor or other health care professional told you that you had gonorrhea, sometimes called GC or clap? Binary variable constructed, 1 if reported yes, 0 if reported no

WSM: We used NHANES[2] to estimate age-specific proportion of women if they were not living with HIV, had >1 female sex partner in the past 12 months, and reported inconsistent condom use or had gonorrhea infection. Proportion with PrEP indications was defined by age group, and we enforced the observed age pattern by defining the PrEP indications in the youngest age group, and the other age groups were defined as a function of the youngest age group.

Sexual activity
- In the past 12 months, with how many men have you had any kind of sex? Binary variable constructed, 1 if reported at least 1, 0 if reported 0

Condom use
- In the past 12 months, about how often have you had vaginal or anal sex without using a condom? Binary variable constructed, 1 if reported 'never' or 'about half the time', 0, if reported 'always', 'less than half the time', or 'not always but more than half the time'

STI
- In the past 12 months, has a doctor or other health care professional told you that you had gonorrhea, sometimes called GC or clap? Binary variable constructed, 1 if reported yes, 0 if reported no.

We checked for missing data in NHANES by variable, based on percentage of people asked the question who did not know or refused to answer. These were excluded from the analysis. The low percentage of non-responses suggests that any non-response is unlikely to have significantly impacted the results. Given the sensitive nature of the sexual behavior questions, there may be biases, such as social desirability bias, impacting the responses.

Level of non-response (does not know or refused to answer) by question:

- In the past 12 months, with how many males have you had anal or oral sex?
    - Less than 1%
- In the past 12 months, with how many men have you had any kind of sex?
    - Less than 1%
- In the past 12 months, with how many men have you had anal sex?
    - Less than 1%
- In the past 12 months, about how often have you had vaginal or anal sex without using a condom?
    - Less than 1%
- In the past 12 months, has a doctor or other health care professional told you that you had chlamydia?
    - Less than 1%

- In the past 12 months, has a doctor or other health care professional told you that you had gonorrhea, sometimes called GC or clap?
    - Less than 1%

PWID: We assumed all PWID are adults aged 18-44. We use published estimates[3] of active PWID and who share needles as having PrEP indications. In the absence of state-specific data, we assumed the proportion of PWID with PrEP indications was the same nationally.

#### 1.2. Risk equation model

##### 1.2.1. HIV acquisition risk through sexual transmission

is modeled as a Bernoulli risk model

$$I_{joar} = 1 - \left[ \left(1 - RR_{or} p_{joa'}\right) + RR_{or} p_{joa'} \left(1 - \left(1 - t_{jo'a}\right)\beta_o\right)^{n_{oa}(1 - s_{oa})} \left(1 - \left(1 - t_{jo'a}\right)\left(1 - d_o\right)\beta_o\right)^{n_{oa} s_{oa}} \right]^{c_{oa}}$$

Where $I_{joar}$ is the estimated annual model-estimated HIV incidence stratified by state (subscript $j$ for US states and Washington DC), sexual transmission population (subscript $o$; MSM, MSW, WSM), age ($a$; age groups modeled 13-17, 18-24, 25-34, 44-54, 55 and older), and race and ethnicity ($r$; populations modeled: Non-Hispanic Black, Hispanic, Non-Hispanic American Indian or Alaska Native [AIAN], Non-Hispanic Native Hawaiian or Pacific Islander [NHOPI], Non-Hispanic Multiracial, and Non-Hispanic Asian).

Below are the variables modeled which consist of behavioral variables, network level variables and epidemiologic variables.
Behavioral factors at the individual level from NHANES, vary by age and sexual behavior but are assumed to be constant across different states. The variables in the Bernoulli model include: $s_{oa}$Probability of using a condom, $c_{oa}$number of partners and $n_{oa}$ the number of acts. We include probability of infection acquisition per act $\beta_o$, which varies by act type: , proportion of acts protected by condom use ($s_{oa}$) and efficacy of condom use ($d_o$).

Network level factors reflect local variation in acquisition risk among people

$RR_{or}$Relative risk of HIV by race/ethnicity and age
$p_{joa'}$ prevalence in the partner pool

The HIV prevalence in the partner pool is a factor in the model and varies by state, sex and sexual behavior, reflecting the differing levels of exposure risk in various demographic groups and geographic locations.

The equation incorporates network-level factors to account for variations in exposure risk among different demographic groups and locations. The prevalence of HIV in the partner pool $p_{joa'}$ varies by age, state, and sexual behavior. This reflects the differing levels of exposure risk across various groups.

Epidemiologic factors included is viral suppression in the partner pool ($t_{jo'a}$), which varies by sexual preference, age, and state. This rate indicates the proportion of people with HIV who maintain an undetectable viral load, thereby reducing the risk of transmission. Additionally, the model considers the acquisition risk by race and ethnicity ($RR_{or}$), acknowledging that different racial and ethnic groups face varying levels of risk. This risk variation by race and ethnicity is influenced by sexual preference but remains constant across states.

##### 1.2.2. HIV acquisition risk through injection drug use

Is modeled as a simpler Bernoulli risk model

$$\boldsymbol{I} = \mathbf{1} - \left[(\mathbf{1} - \boldsymbol{p_j}) + \boldsymbol{p_j}(\mathbf{1} - (\mathbf{1} - \boldsymbol{t_j})\boldsymbol{\beta})^{rn/c}\right]^{c}$$

With I the average HIV incidence among PWID

$\boldsymbol{p_j}$ is HIV prevalence among PWID in the state, which we approximate by calculating 3-year cumulative diagnoses among PWID in the state. This is done to account for rapid turnover[23] in the population which may mean not all people who acquired HIV via IDU are still active PWID. Given the small population size we do not attempt to examine HIV prevalence by sex, race/ethnicity or age within states.

$\boldsymbol{t_j}$ is the viral suppression among PWID living with HIV

$\boldsymbol{n}$ is the annual number of injections, $\boldsymbol{c}$ is the annual number of needle-sharing partners, and $\boldsymbol{r}$ is the proportion of injections which include receptive needle-sharing

$\boldsymbol{\beta}$ is the HIV acquisition probability per receptive needle-sharing act

#### 1.3. Calibration

Sampling Importance Resampling (SIR)
To calibrate the state-level model to national-level data, we used Sampling Importance Resampling (SIR). Initial Sampling: Parameters were sampled from their respective prior distributions. National-level priors were used by age and transmission risk group, but constant across states. We calculated likelihood for each draw based on calibration targets. We then resampled parameter sets with replacements based on the importance weights. Importance weights were calculated based on proportional likelihood.

**Table S1.** Parameters used in the risk model, which were applied at the national level (assumed to be constant between states)

| Parameter | Prior distribution | Shape parameters | Median (95%UI) of the prior distribution | Reference |
|---|---|---|---|---|
| Sexual acquisition risk model parameters | | | | |
| Proportion of MSM with PrEP indicators | | | | [2] |
| MSM 18-44 yos | Beta | (1552.69, 858.81) | 0.644 (0.63-0.66 ) | |
| MSM 45-54, 55+ yos | Beta | (499.11, 350.37) | 0.588 (0.55-0.62) | |
| Proportion of women with PrEP indicators | | | | [2] |
| WSM 18-24 yos (reference) | Beta | (167.98, 521.08) | 0.244 (0.213-0.276) | |
| WSM 25-34 yos (RR) | Fixed | - | 0.58 | |
| WSM 35-44 yos (RR) | Fixed | - | 0.39 | |
| WSM 45-54 yos (RR) | Fixed | - | 0.21 | |
| WSM 55+ yos (RR) | Fixed | - | 0.17 | |
| Proportion of MSW with PrEP indicators | | | | [2] |
| MSW 18-24 yos (reference) | Beta | (124.74, 358.55) | 0.258 (0.220-0.298) | |
| MSW 25-34 yos (RR) | Fixed | - | 0.65 | |
| MSW 35-44 yos (RR) | Fixed | - | 0.44 | |
| MSW 45-54 yos (RR) | Fixed | - | 0.34 | |
| MSW 55+ yos (RR) | Fixed | - | 0.17 | |
| Proportion of sexually active 13-17 yos with PrEP indicators (MSM, MSW, WSM) | Beta | (8.26, 30.87) | 0.207 (0.10-0.35) | [4,5] |
| Relative rate of HIV prevalence in partner pool* | | | | [6] |
| MSM | | | | |
| White (reference) | Fixed | | 1 | |
| RR Black | Uniform | (4.14, 5.06) | 4.6 (4.16, 5.03) | |
| RR Hispanic | Uniform | (2.17, 2.66) | 2.42 (2.19, 2.65) | |
| RR AIAN | Uniform | (0.834, 1.02) | 0.926 (0.838, 1.01) | |
| RR NHOPI | Uniform | (1.33, 1.62) | 1.48 (1.34, 1.62) | |
| RR Multiracial | Uniform | (0.9, 1.1) | 1 (0.905, 1.1) | |
| RR Asian | Uniform | (0.726, 0.887) | 0.806 (0.73, 0.883) | |
| WSM | | | | |
| White (reference) | Fixed | | 1 | |
| RR Black | Uniform | (18.4, 22.5) | 20.4 (18.5, 22.4) | |
| RR Hispanic | Uniform | (3.7, 4.52) | 4.11 (3.72, 4.5) | |
| RR AIAN | Uniform | (1.9, 2.32) | 2.11 (1.91, 2.31) | |
| RR NHOPI | Uniform | (2.4, 2.93) | 2.67 (2.41, 2.92) | |
| RR Multiracial | Uniform | (1, 1.22) | 1.11 (1.01, 1.22) | |
| RR Asian | Uniform | (0.9, 1.1) | 1 (0.905, 1.1) | |

| | | | | |
|---|---|---|---|---|
| MSW | | | | |
| White (reference) | | | | |
| RR Black | Uniform | (23.2, 28.3) | 25.8 (23.3, 28.2) | |
| RR Hispanic | Uniform | (5.18, 6.32) | 5.75 (5.2, 6.3) | |
| RR AIAN | Uniform | (2.02, 2.48) | 2.25 (2.04, 2.46) | |
| RR NHOPI | Uniform | (0.675, 0.825) | 0.75 (0.679, 0.821) | |
| RR Multiracial | Uniform | (1.12, 1.38) | 1.25 (1.13, 1.37) | |
| RR Asian | Uniform | (0.9, 1.1) | 1 (0.905, 1.1) | |
| MSM, transmission probability per act (AI average of insertive and receptive) | Beta | (37.4, 4960) | 0.00741 (0.00528, 0.01) | [7] |
| MSM number of acts per partnership | Log-normal | (2.538,0.587) | 12.68 (4.01, 39.63) | Assumption |
| MSM number of partners per year | | | | [2] |
| 13-24 yos | Log-normal | (1.93, 0.24) | 3.21 (2.01-5.12) | |
| 25-34 yos | Log-normal | (1.93, 0.24) | 3.21 (2.01-5.12) | |
| 35-44 yos | Log-normal | (1.93, 0.24) | 3.21 (2.01-5.12) | |
| 45-54 yos | Log-normal | (0.49, 0.18) | 1.63 (1.15-2.30) | |
| 55+ yos | Log-normal | (0.49, 0.18) | 1.63 (1.15-2.30) | |
| MSM condom use in partnerships (proportion of acts) | | | | [2] |
| 13-24 yos | Beta | (7.99, 24.4) | 0.242 (0.117, 0.406) | |
| 25-34 yos | Beta | (7.99, 24.4) | 0.242 (0.117, 0.406) | |
| 35-44 yos | Beta | (7.99, 24.4) | 0.242 (0.117, 0.406) | |
| 45-54 yos | Beta | (7.99, 24.4) | 0.242 (0.117, 0.406) | |
| 55+ yos | Beta | (7.66, 16.9) | 0.307 (0.149, 0.505) | |
| Condom effectiveness in MSM | Beta | (56.6, 24.4) | 0.7 (0.595, 0.793) | [8] |
| WSM transmission probability per act (VI receptive) | Beta | (12.8, 10000) | 0.00125 (0.000679, 0.00207) | [9] |
| WSM number of acts per partner | Log-normal | (2.538,0.587) | 12.68 (4.01, 39.63) | Assumption |
| WSM number of partners per year | | | | [2] |
| 13-24 yos | Log-normal | (1.5, 0.155) | 4.47 (3.3, 6.05) | |
| 25-34 yos | Log-normal | (1.38, 0.095) | 3.99 (3.31, 4.8) | |
| 35-44 yos | Log-normal | (1.34, 0.132) | 3.84 (2.96, 4.97) | |
| 45-54 yos | Log-normal | (1.6, 0.184) | 4.95 (3.45, 7.1) | |
| 55+ yos | Log-normal | (1.67, 0.231) | 5.32 (3.38, 8.36) | |
| WSM condom use in partnerships (proportion of acts) | | | | [2] |
| 13-24 yos | Beta | (104, 295) | 0.261 (0.22, 0.306) | |
| 25-34 yos | Beta | (105, 375) | 0.219 (0.183, 0.257) | |
| 35-44 yos | Beta | (51.1, 211) | 0.194 (0.15, 0.245) | |
| 45-54 yos | Beta | (15.9, 119) | 0.116 (0.069, 0.177) | |

| | | | | |
|---|---|---|---|---|
| 55+ yos | Beta | (1.11, 40.8) | 0.0195 (0.000947, 0.0916) | |
| Condom effectiveness in WSM | Beta | (53.3, 13.5) | 0.8 (0.694, 0.884) | [10] |
| MSW transmission probability per act (VI insertive) | Beta | (2.97, 6590) | 0.000401 (9.19e-05, 0.00109) | [9] |
| MSW number of acts per partnership | Log-normal | (2.538,0.587) | 12.68 (4.01, 39.63) | Assumption |
| MSW number of partners per year | | | | [2] |
| 13-24 yos | Log-normal | (1.98, 0.255) | 7.28 (4.42, 12) | |
| 25-34 yos | Log-normal | (1.54, 0.077) | 4.66 (4.01, 5.42) | |
| 35-44 yos | Log-normal | (1.58, 0.133) | 4.84 (3.73, 6.28) | |
| 45-54 yos | Log-normal | (1.51, 0.145) | 4.51 (3.39, 5.99) | |
| 55+ yos | Log-normal | (1.7, 0.118) | 5.46 (4.33, 6.88) | |
| MSW condom use in partnerships (proportion of acts) | | | | [2] |
| 13-24 yos | Beta | (166, 416) | 0.285 (0.249, 0.322) | |
| 25-34 yos | Beta | (127, 359) | 0.261 (0.223, 0.301) | |
| 35-44 yos | Beta | (54.7, 203) | 0.212 (0.165, 0.264) | |
| 45-54 yos | Beta | (48.9, 193) | 0.202 (0.154, 0.255) | |
| 55+ yos | Beta | (14.1, 50.3) | 0.216 (0.128, 0.327) | |
| Condom effectiveness in MSW | Beta | (53.3, 13.5) | 0.8 (0.694, 0.884) | [10] |
| PWID model parameters | | | | |
| Proportion actively injecting pwid, men | Beta | (7.08, 330) | 0.0201 (0.00856, 0.0388) | [3] |
| Proportion actively injecting pwid, women | Beta | (7.77, 838) | 0.0088 (0.00392, 0.0166) | [3] |
| Average number of injections annually, men | Normal | (569, 77.3) | 569 (417, 720) | [2] |
| Average number of injections annually, women | Normal | (867, 169) | 867 (536, 1200) | [2] |
| Number of injection sharing partners (same for both sexes) | Gamma | (10, 2.43) | 4 (1.98, 7.05) | [11] |
| Proportion of receptive needle-sharing injections, men | Beta | (40.9, 29) | 0.586 (0.469, 0.697) | [12] |
| Proportion of receptive needle-sharing injections, women | Beta | (36.8, 22) | 0.627 (0.5, 0.744) | [12] |
| HIV transmission probability per needle-share act | Beta | (0.69, 1000) | 0.000398 (4.15e-06, 0.00299) | [13] |

MSM: Men who have sex with men; MSW: men who have sex with women; RR: relative rate; UI: uncertainty interval; WSM: women who have sex with men
* All groups beside Hispanic are non-Hispanic (groups are mutually exclusive)

#### 2. **Cost-effectiveness threshold approach**

Willingness to pay approach starts by estimating national level of willingness to pay for PrEP that is then allocated based on need (top-down approach)

##### 2.1. **Equation**

The cost-effectiveness threshold approach incorporates cost considerations into the calculation of PrEP need. This involves estimating how many people should be treated to avert one HIV infection, given a willingness to pay (WTP) threshold of $100,000 per quality-adjusted life year (QALY).

$$\frac{Annual\ PrEP\ cost\ * X - cost\ of\ HIV\ infection}{QALYs\ per\ infection} = \$100{,}000$$

The number of people (X) required is calculated using the equation:

$$X = \frac{\$100{,}000 * QALYs\ per\ infection + cost\ of\ HIV\ infection}{Annual\ PrEP\ cost}$$

##### 2.2. **Data**

The calculation employed the annual cost of PrEP use ($12,376),[14,15] lifetime costs associated with HIV infection ($478,142),[14,16] and 4.45 QALYs lost per HIV infection.[14,16] This corresponds to 75 persons using PrEP to avert one infection given a willingness to pay (WTP) 100,000 per QALY gained.

**Table S2.** HIV incidence estimates used in the analyses (CDC 2022)[17]

| **Sex** | **Transmission Category** | **Incidence** | **PrEP need based on WTP** |
|---|---|---|---|
| Male | MSM | 21,100 | 1,582,500 |
| All | PWID | 2,500 | 187,500 |
| Female | PHET - women | 5,100 | 382,500 |
| Male | PHET - men | 2,000 | 150,000 |
| All | All | 30,700 | 2,302,500 |

MSM: Men who have sex with men; MSW: men who have sex with women; RR: relative rate; UI: uncertainty interval; WSM: women who have sex with men; WTP: willingness to pay

### 3. Results

#### 3.1. Calibration results

**Figure S2.** Calibration to national-level incidence

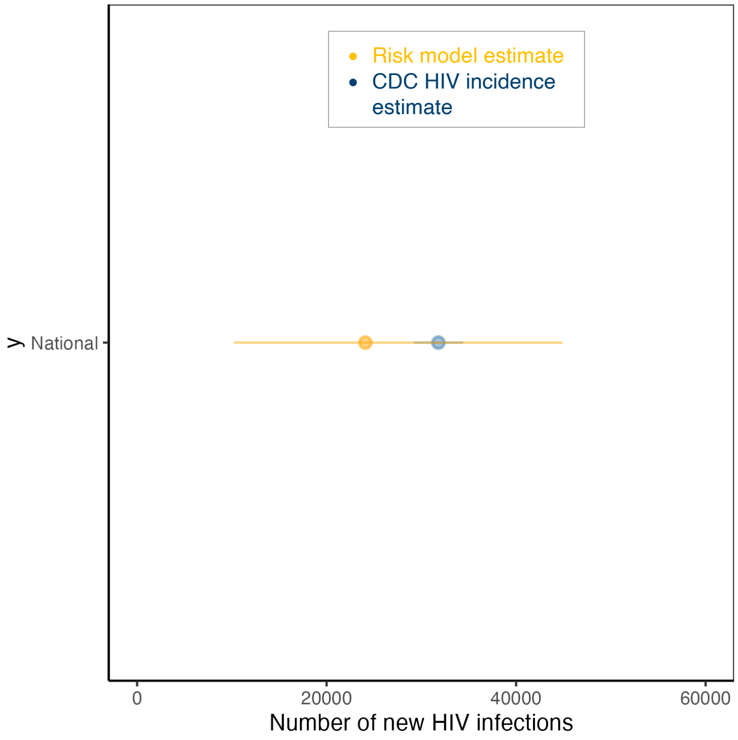

**Figure S3.** Calibration to transmission risk group of new HIV infections.

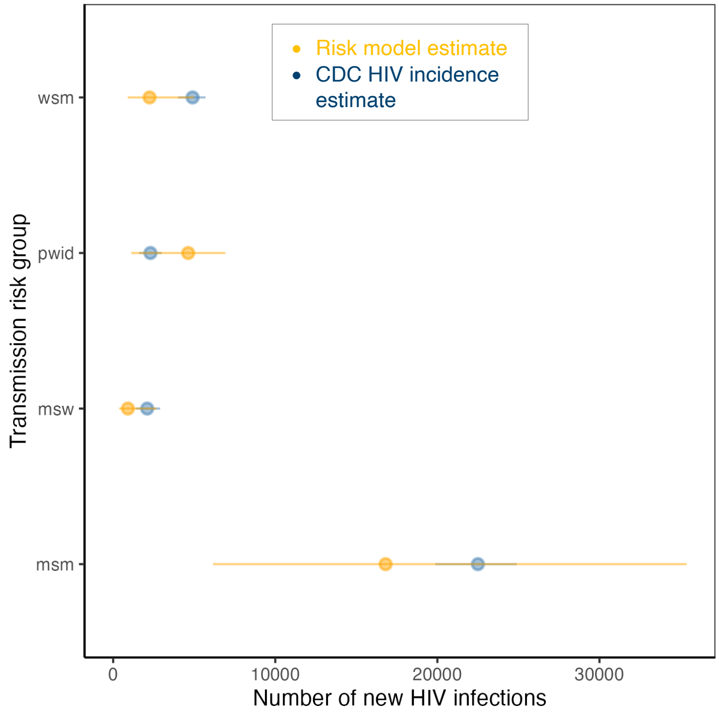

**Figure S4.** Calibration to age distribution of new HIV infections across populations

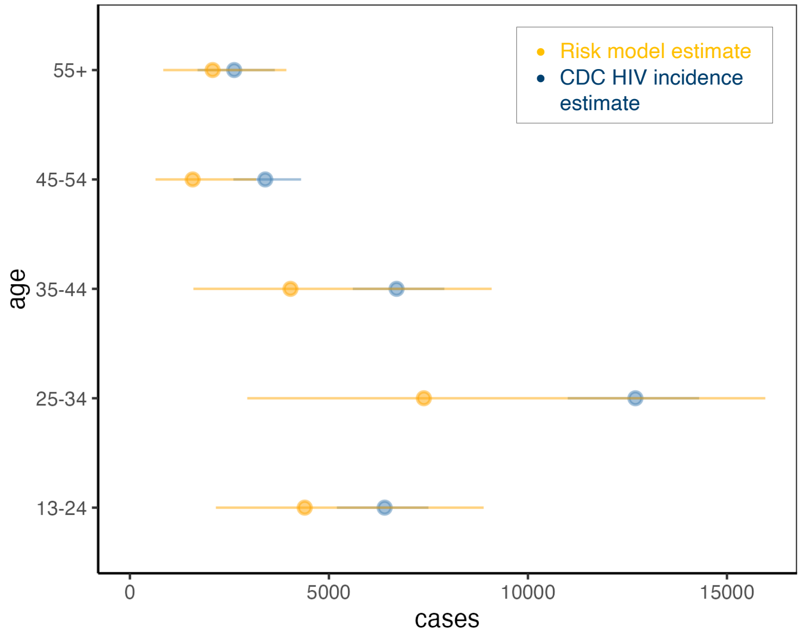

**Figure S5.** Calibration to age distribution of new HIV infections in MSM

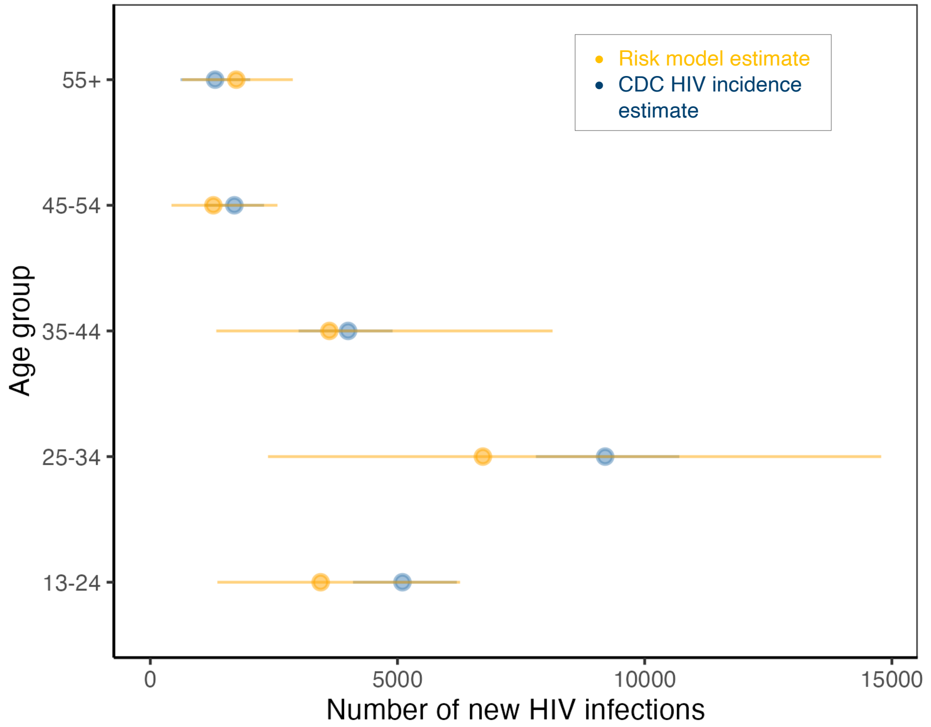

**Figure S6.** Calibration to race/ethnicity distribution of new HIV infections

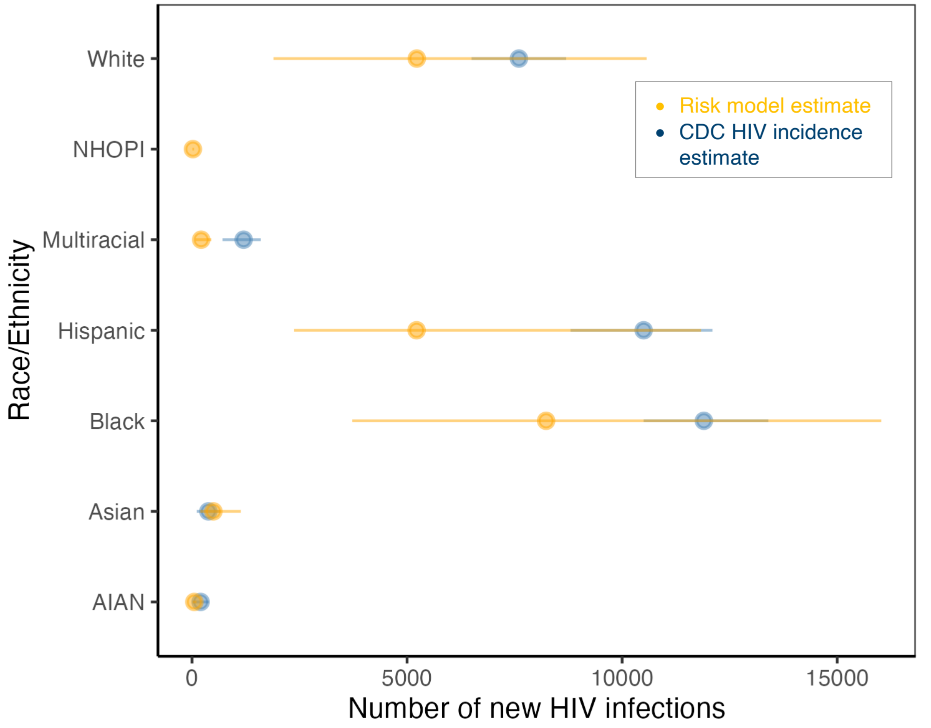

**Figure S7.** Calibration to sex among PWID of new HIV infections

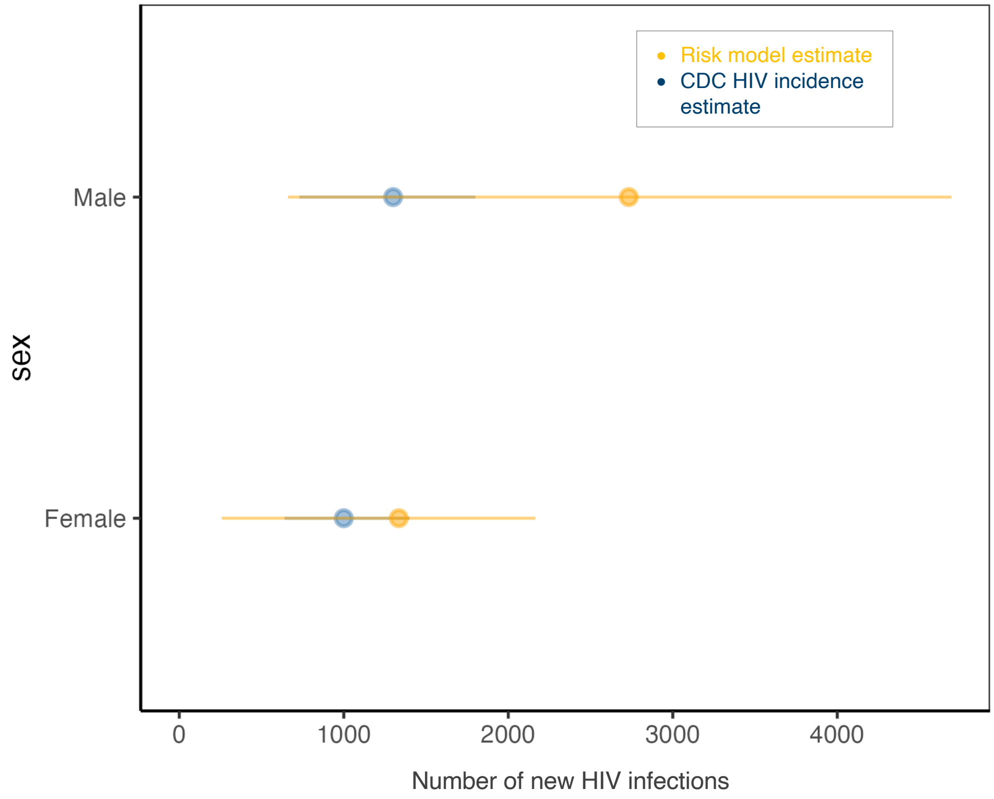

**Figure S8.** Calibration to state-level incidence. Panel on the left showcases states which had incidence estimates available, and the panel on the right showcases states for which incidence estimates were not available, model estimated HIV incidence is shown.

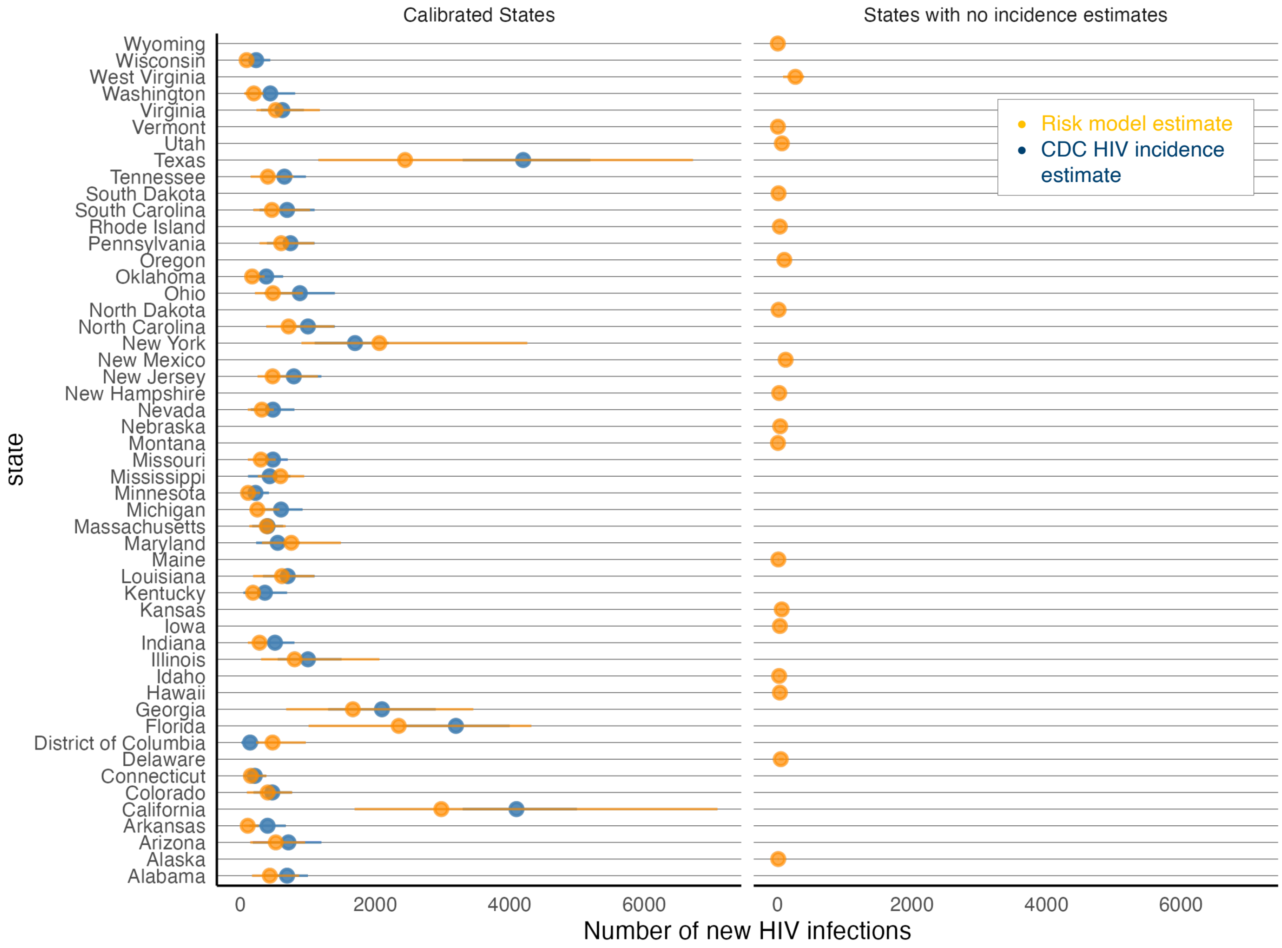

##### 3.2. HIV risk model estimates by transmission risk group

**Figure S9.** Panel A shows the cumulative proportion (y-axis) of number of new HIV infections within a transmission risk group by the average annual HIV acquisition risk per 100 persons (x-axis). Panel B shows the cumulative proportion of the population size by the average annual HIV acquisition risk per 100 persons (x-axis). The x axes vary between the different populations to reflect the varying HIV risk. Panel C compares these two cumulative measures by using a Lorenz curve (red line represents equality). Each line represents a simulation draw from the calibrated model.

A) HIV infections by HIV risk

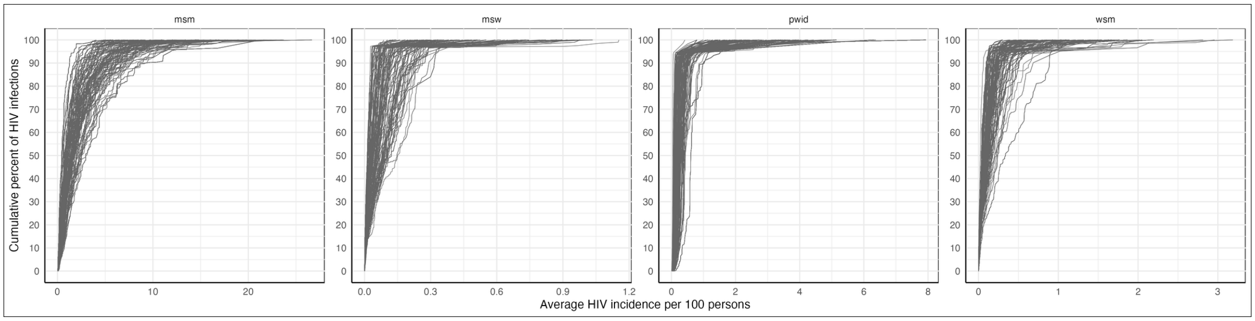

B) Population size by HIV risk

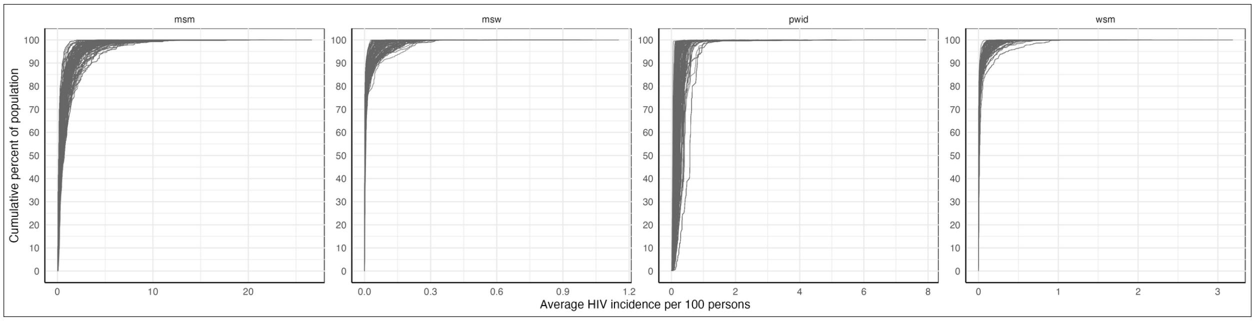

C) Lorenz curve of population size and HIV infections

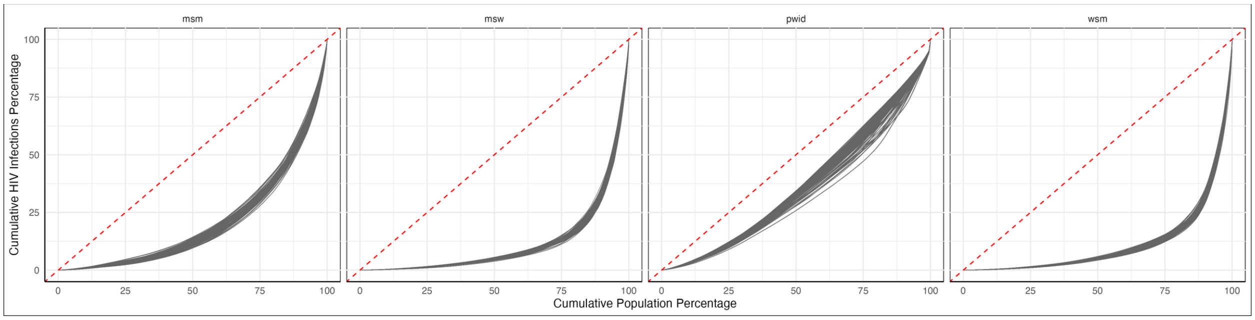

##### 3.3. Tabulations of number who could benefit from PrEP by allocation criteria

**Table S3.** Estimates of need for PrEP by transmission risk group, total number

| Transmission Risk Group | Former CDC estimate [a] | Cost-effectiveness threshold estimate [b] | HIV acquisition risk thresholds [c] | | |
|---|---|---|---|---|---|
| | | | 100% of HIV infections | 90% of HIV infections | 75% of HIV infections |
| **MSM** | 1,180,000 (NA-NA) | 1,580,000 (NA-NA) | 2,970,000 (2,900,000-3,060,000) | 2,420,000 (2,170,000-2,700,000) | 1,620,000 (1,380,000-1,890,000) |
| **MSW** | 111,000 (NA-NA) | 150,000 (NA-NA) | 13,400,000 (11,400,000-15,300,000) | 310,000 (3,800-844,000) | 13,500 (0-74,300) |
| **PWID** | 136,000 (NA-NA) | 188,000 (NA-NA) | 1,600,000 (1,120,000-2,100,000) | 1,410,000 (695,000-2,060,000) | 708,000 (8,310-1,420,000) |
| **WSM** | 252,000 (NA-NA) | 382,000 (NA-NA) | 12,100,000 (11,300,000-13,700,000) | 622,000 (141,000-1,300,000) | 99,600 (6,590-529,000) |

MSM: Men who have sex with men; MSW: men who have sex with women; PWID: people who inject drugs; WSM: women who have sex with men

a) Relying on HIV diagnosis data reported in 2022
b) Relying on published estimates in the literature
c) Relying on NHANES estimates (2013-2016), published estimates in the literature, and 2022 incidence estimates

**Table S4.** Estimates of need for PrEP by transmission risk population, per 100 person

| Transmission Risk Group | Former CDC estimate | Cost-effectiveness threshold estimate | HIV acquisition risk thresholds | | |
|---|---|---|---|---|---|
| | | | 100% of HIV infections | 90% of HIV infections | 75% of HIV infections |
| **MSM** | 25 (NA-NA) | 30 (NA-NA) | 57 (56-59) | 47 (42-52) | 31 (27-36) |
| **MSW** | 0 (NA-NA) | 0 (NA-NA) | 11 (9-12) | 0 (0-1) | 0 (0-0) |
| **PWID** | 5 (NA-NA) | 7 (NA-NA) | 60 (52-66) | 53 (28-61) | 26 (0-44) |
| **WSM** | 0 (NA-NA) | 0 (NA-NA) | 9 (8-10) | 0 (0-1) | 0 (0-0) |

MSM: Men who have sex with men; MSW: men who have sex with women; PWID: people who inject drugs; WSM: women who have sex with men

**Table S5.** HIV acquisition risk estimates of need for PrEP by threshold and transmission risk group and age, total number

| Transmission Risk Group | Age | 100% of HIV infections | 90% of HIV infections | 75% of HIV infections |
|---|---|---|---|---|
| **MSM** | 13-17 | 52,700 (30,700-69,800) | 52,200 (30,600-67,500) | 44,400 (27,400-62,100) |
| | 18-24 | 355,000 (339,000-371,000) | 352,000 (337,000-371,000) | 298,000 (257,000-341,000) |
| | 25-34 | 579,000 (566,000-589,000) | 579,000 (566,000-589,000) | 540,000 (519,000-581,000) |
| | 35-44 | 516,000 (504,000-525,000) | 510,000 (494,000-523,000) | 419,000 (355,000-487,000) |
| | 45-54 | 473,000 (450,000-499,000) | 361,000 (270,000-446,000) | 152,000 (61,800-270,000) |
| | 55+ | 963,000 (914,000-1,010,000) | 533,000 (359,000-772,000) | 143,000 (65,600-268,000) |
| **MSW** | 13-17 | 513,000 (310,000-757,000) | 20,500 (296-43,600) | 953 (0-7,160) |
| | 18-24 | 2,200,000 (1,780,000-3,050,000) | 87,100 (1,410-186,000) | 3,960 (0-34,200) |
| | 25-34 | 3,560,000 (3,040,000-4,080,000) | 66,900 (0-240,000) | 2,410 (0-3,610) |
| | 35-44 | 2,170,000 (1,850,000-2,480,000) | 27,500 (0-109,000) | 1,290 (0-1,850) |
| | 45-54 | 1,700,000 (1,450,000-1,950,000) | 20,000 (0-83,700) | 872 (0-1,400) |
| | 55+ | 1,750,000 (1,490,000-2,000,000) | 29,100 (0-104,000) | 1,250 (0-10,100) |
| **WSM** | 13-17 | 482,000 (283,000-685,000) | 26,300 (4,180-57,100) | 4,160 (113-27,000) |
| | 18-24 | 1,990,000 (1,450,000-3,170,000) | 108,000 (17,700-249,000) | 15,300 (376-81,400) |
| | 25-34 | 3,080,000 (2,930,000-3,440,000) | 147,000 (21,900-322,000) | 19,600 (0-91,700) |
| | 35-44 | 1,920,000 (1,830,000-2,150,000) | 81,800 (15,000-195,000) | 9,550 (0-54,800) |
| | 45-54 | 1,070,000 (1,020,000-1,200,000) | 75,600 (22,800-129,000) | 15,900 (957-66,700) |

| | | | | |
|---|---|---|---|---|
| | 55+ | 2,030,000 (1,930,000-2,270,000) | 99,900 (37,500-198,000) | 22,000 (2,060-83,900) |

MSM: Men who have sex with men; MSW: men who have sex with women; PWID: people who inject drugs; WSM: women who have sex with men

**Table S6.** HIV acquisition risk estimates of need for PrEP by threshold and transmission risk group and age, per 100 person

| Transmission Risk Group | Age | 100% of HIV infections | 90% of HIV infections | 75% of HIV infections |
|---|---|---|---|---|
| **MSM** | 13-17 | 9 (5-12) | 9 (5-11) | 7 (5-10) |
| | 18-24 | 35 (33-36) | 34 (33-36) | 29 (25-33) |
| | 25-34 | 64 (63-65) | 64 (63-65) | 60 (58-64) |
| | 35-44 | 64 (63-65) | 63 (62-65) | 52 (44-61) |
| | 45-54 | 58 (55-62) | 44 (33-55) | 19 (8-33) |
| | 55+ | 58 (55-62) | 32 (22-47) | 9 (4-16) |
| **MSW** | 13-17 | 3 (2-5) | 0 (0-0) | 0 (0-0) |
| | 18-24 | 9 (7-12) | 0 (0-1) | 0 (0-0) |
| | 25-34 | 16 (14-19) | 0 (0-1) | 0 (0-0) |
| | 35-44 | 11 (9-13) | 0 (0-1) | 0 (0-0) |
| | 45-54 | 9 (7-10) | 0 (0-0) | 0 (0-0) |
| | 55+ | 4 (4-5) | 0 (0-0) | 0 (0-0) |
| **WSM** | 13-17 | 3 (2-5) | 0 (0-0) | 0 (0-0) |
| | 18-24 | 8 (6-13) | 0 (0-1) | 0 (0-0) |
| | 25-34 | 14 (13-16) | 1 (0-1) | 0 (0-0) |
| | 35-44 | 9 (9-10) | 0 (0-1) | 0 (0-0) |
| | 45-54 | 5 (5-6) | 0 (0-1) | 0 (0-0) |
| | 55+ | 4 (4-5) | 0 (0-0) | 0 (0-0) |

MSM: Men who have sex with men; MSW: men who have sex with women; PWID: people who inject drugs; WSM: women who have sex with men

**Table S7.** HIV acquisition risk estimates of need for PrEP by threshold and transmission risk group and race/ethnicity, total number

| Transmission Risk Group | Race/Ethnicity | 100% of HIV infections | 90% of HIV infections | 75% of HIV infections |
|---|---|---|---|---|
| **MSM** | AIAN | 16,400 (16,000-16,900) | 12,300 (10,100-14,300) | 6,540 (3,560-8,600) |
| | Asian | 187,000 (183,000-192,000) | 139,000 (113,000-170,000) | 76,700 (55,200-101,000) |
| | Black | 336,000 (329,000-347,000) | 336,000 (329,000-347,000) | 322,000 (302,000-338,000) |
| | Hispanic | 573,000 (561,000-590,000) | 569,000 (560,000-589,000) | 486,000 (394,000-554,000) |
| | Multiracial | 56,600 (55,300-58,400) | 48,400 (43,400-53,100) | 32,000 (23,000-38,300) |
| | NHOPI | 5,000 (4,900-5,160) | 4,420 (3,980-4,880) | 2,910 (2,040-3,730) |
| | White | 1,800,000 (1,750,000-1,850,000) | 1,310,000 (1,090,000-1,550,000) | 694,000 (601,000-872,000) |
| **MSW** | AIAN | 91,600 (77,500-104,000) | 0 (0-0) | 0 (0-0) |
| | Asian | 755,000 (640,000-863,000) | 0 (0-0) | 0 (0-0) |
| | Black | 1,680,000 (1,430,000-1,920,000) | 309,000 (3,800-840,000) | 13,500 (0-74,300) |
| | Hispanic | 2,620,000 (2,220,000-2,990,000) | 903 (0-3,590) | 0 (0-0) |
| | Multiracial | 321,000 (271,000-365,000) | 0 (0-0) | 0 (0-0) |
| | NHOPI | 24,200 (20,500-27,600) | 0 (0-0) | 0 (0-0) |
| | White | 7,920,000 (6,710,000-9,060,000) | 0 (0-0) | 0 (0-0) |
| **WSM** | AIAN | 83,500 (78,000-94,600) | 12 (0-72) | 0 (0-0) |
| | Asian | 737,000 (690,000-832,000) | 4 (0-79) | 0 (0-0) |
| | Black | 1,610,000 (1,500,000-1,820,000) | 612,000 (141,000-1,250,000) | 99,400 (6,590-526,000) |
| | Hispanic | 2,270,000 (2,120,000-2,580,000) | 9,740 (0-60,200) | 229 (0-2,600) |
| | Multiracial | 297,000 (275,000-337,000) | 4 (0-45) | 0 (0-0) |
| | NHOPI | 21,800 (20,300-24,600) | 3 (0-13) | 0 (0-0) |
| | White | 7,090,000 (6,640,000-8,010,000) | 43 (0-622) | 0 (0-0) |

AIAN: American Indian or Alaska Native; MSM: Men who have sex with men; MSW: men who have sex with women; NHOPI: Native Hawaiian or other Pacific Islander; PWID: people who inject drugs; WSM: women who have sex with men

**Table S8.** HIV acquisition risk estimates of need for PrEP by threshold and transmission risk group and race/ethnicity, per 100 person

| Transmission Risk Group | Race/Ethnicity | 100% of HIV infections | 90% of HIV infections | 75% of HIV infections |
|---|---|---|---|---|
| **MSM** | AIAN | 57 (56-59) | 43 (35-49) | 23 (12-30) |
| | Asian | 58 (57-60) | 43 (35-53) | 24 (17-31) |
| | Black | 57 (56-59) | 57 (56-59) | 54 (51-57) |
| | Hispanic | 57 (55-58) | 56 (55-58) | 48 (39-55) |
| | Multiracial | 55 (54-57) | 47 (42-52) | 31 (22-37) |
| | NHOPI | 57 (56-59) | 51 (45-56) | 33 (23-43) |
| | White | 58 (56-59) | 42 (35-50) | 22 (19-28) |
| **MSW** | AIAN | 11 (9-13) | 0 (0-0) | 0 (0-0) |
| | Asian | 11 (9-13) | 0 (0-0) | 0 (0-0) |
| | Black | 11 (10-13) | 2 (0-6) | 0 (0-0) |
| | Hispanic | 12 (10-14) | 0 (0-0) | 0 (0-0) |
| | Multiracial | 13 (11-15) | 0 (0-0) | 0 (0-0) |
| | NHOPI | 12 (10-13) | 0 (0-0) | 0 (0-0) |
| | White | 10 (8-11) | 0 (0-0) | 0 (0-0) |
| **WSM** | AIAN | 9 (9-10) | 0 (0-0) | 0 (0-0) |
| | Asian | 9 (9-10) | 0 (0-0) | 0 (0-0) |
| | Black | 9 (9-10) | 4 (1-7) | 1 (0-3) |
| | Hispanic | 10 (9-12) | 0 (0-0) | 0 (0-0) |
| | Multiracial | 11 (10-13) | 0 (0-0) | 0 (0-0) |
| | NHOPI | 10 (9-11) | 0 (0-0) | 0 (0-0) |
| | White | 8 (8-9) | 0 (0-0) | 0 (0-0) |

AIAN: American Indian or Alaska Native; MSM: Men who have sex with men; MSW: men who have sex with women; NHOPI: Native Hawaiian or other Pacific Islander; PWID: people who inject drugs; WSM: women who have sex with men

#### 3.4. State-level results

**Figure S10.** People who could benefit from PrEP based on different criteria used. Panel A displays the number of people by state (x-axis is in log scale), and Panel B displays the percentage of people who could benefit from PrEP per total population 13 and older.

A) Number of people who could benefit from PrEP, ordered by number to benefit from PrEP

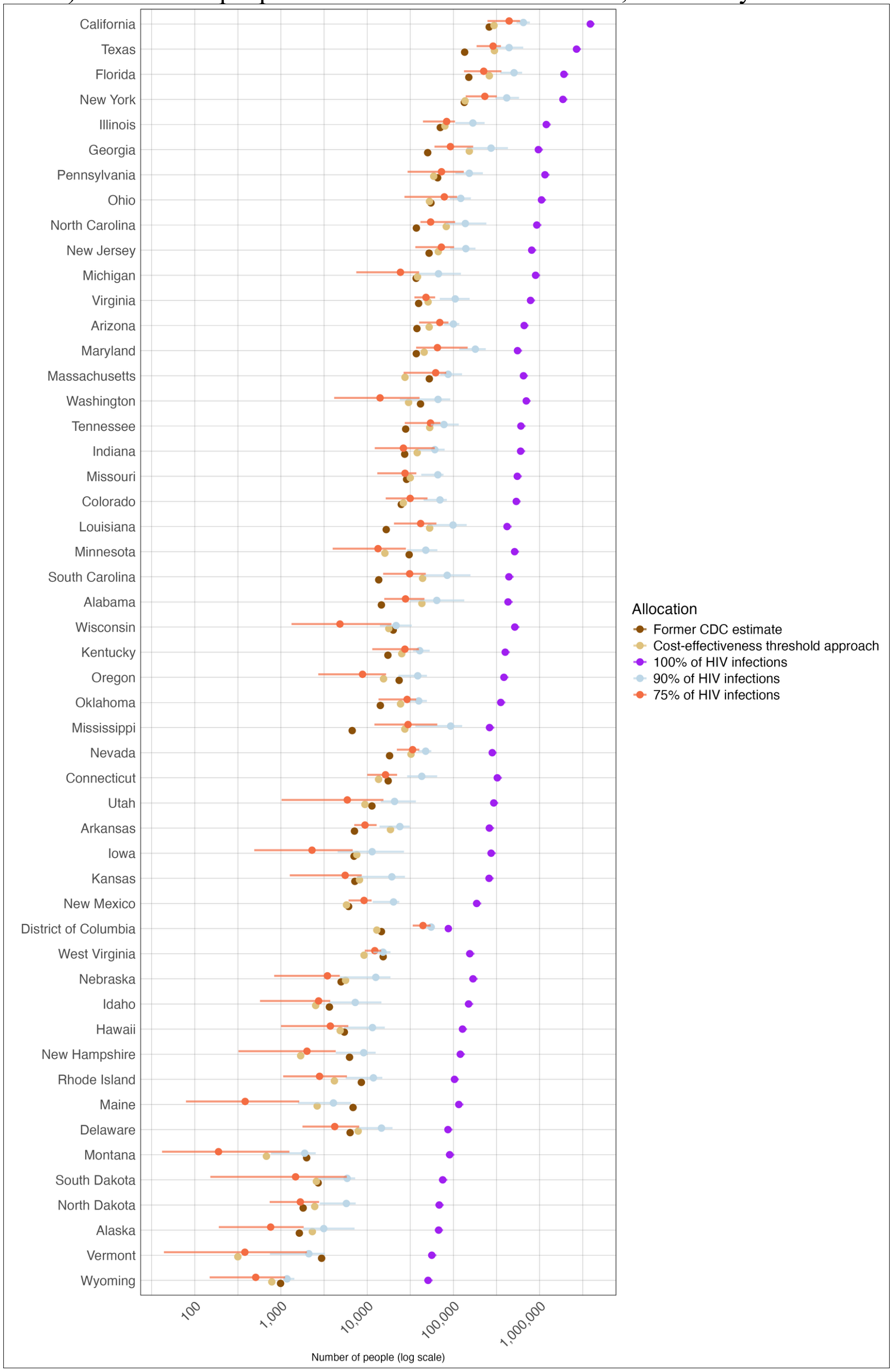

B) Rate of people who could benefit from PrEP per total population 13 and older

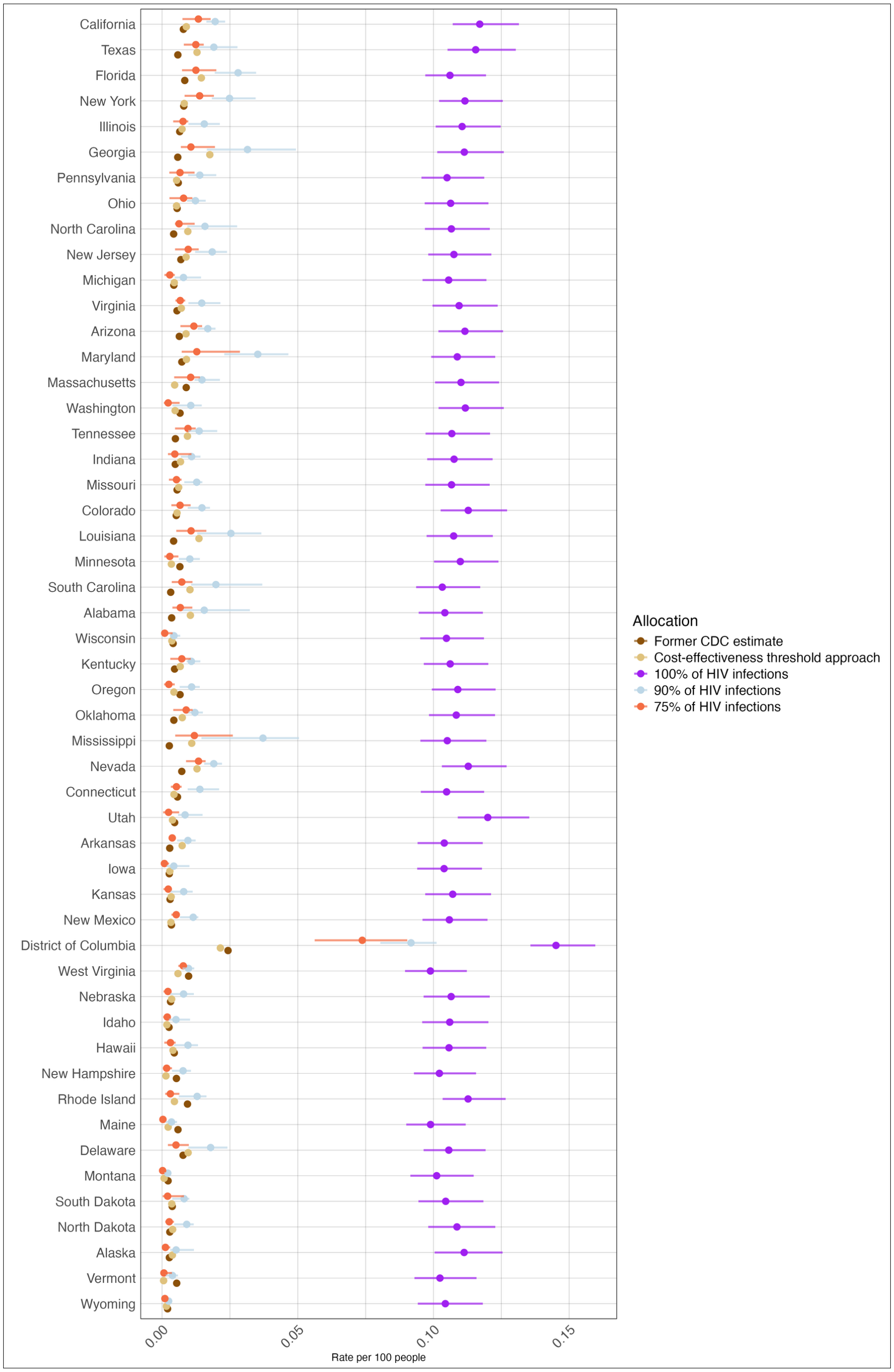
